# High-resolution epigenome analysis in nasal samples derived from children with respiratory viral infections reveals striking changes upon SARS-CoV-2 infection

**DOI:** 10.1101/2021.03.09.21253155

**Authors:** Konner Winkley, Boryana Koseva, Dithi Banerjee, Warren Cheung, Rangaraj Selvarangan, Tomi Pastinen, Elin Grundberg

## Abstract

**Background:** DNA methylation patterns of the human genome can be modified by environmental stimuli and provide dense information on gene regulatory circuitries. We studied genome-wide DNA methylation in nasal samples from infants (<6 months) applying whole-genome bisulfite sequencing (WGBS) to characterize epigenome response to 10 different respiratory viral infections including SARS-CoV-2.

**Results:** We identified virus-specific differentially methylated regions (vDMR) with human metapneumovirus (hMPV) and SARS-CoV-2 followed by Influenza B (Flu B) causing the weakest vs. strongest epigenome response with 496 vs. 78541 and 14361 vDMR, respectively. We found a strong replication rate of FluB (52%) and SARS-CoV-2 (42%) vDMR in independent samples indicating robust epigenome perturbation upon infection. Among the FluB and SARS-CoV-2 vDMRs, around 70% were hypomethylated and significantly enriched among epithelial cell-specific regulatory elements whereas the hypermethylated vDMRs for these viruses mapped more frequently to immune cell regulatory elements, especially those of the myeloid lineage. The hypermethylated vDMRs were also enriched among genes and genetic loci in monocyte activation pathways and monocyte count. Finally, we perform single-cell RNA-sequencing characterization of nasal mucosa in response to these two viruses to functionally analyze the epigenome perturbations. Which supports the trends we identified in methylation data and highlights and important role for monocytes.

**Conclusions:** All together, we find evidence indicating genetic predisposition to innate immune response upon a respiratory viral infection. Our genome-wide monitoring of infant viral response provides first catalogue of associated host regulatory elements. Assessing epigenetic variation in individual patients may reveal evidence for viral triggers of childhood disease.

## Background

Methylation of cytosine bases in the CpG context plays an important role in controlling gene expression and can be modified by environmental and biological stimuli. This has led to methylation patterns being studied in many contexts including complex disease (Allum et al. 2019; Liang et al. 2015), infection (Maeda et al. 2017; Matsusaka et al. 2017; Mcerlean et al. 2014), and developmental processes (Smith et al. 2012; Okano et al. 1999). A key analysis in these studies is the identification of which genomic regions change in methylation level in response to the stimulus of interest. We and others have shown that variable and disease-associated DNA methylation regions map to regulatory regions, specifically enhancers (Grundberg et al. 2013; Allum et al. 2015). These differentially methylated regions (DMRs) provide information about pathways that may be activated or suppressed in response to environmental stimuli (Busche et al. 2015; Tsaprouni et al. 2014). Because of this, whole genome bisulfite sequencing (WGBS) - the gold standard method for assessing genome-wide DNA methylation at single base resolution - can provide genome-scale insight into the mechanism of action for biological phenomenon.

The COVID-19 pandemic has led to a dramatic surge in research on the host response mechanisms to respiratory virus infection, specifically in response to SARS-CoV-2 infection. However, there are several other respiratory viruses beyond SARS-CoV-2 that are known to cause acute respiratory illness (ARI), and there are still gaps in knowledge about the interplay between the host immune system and viral replication in many if not all of these viruses. While these viruses are evolutionarily distant (Stec et al. 1991; Collins et al. 2013; Lu et al. 2020), the clinical presentation of illness from their infection is quite similar. It is therefore unknown if these viruses are more similar to, or more divergent from one another in the host immune responses they illicit upon infection. Additionally, there are viruses such as Respiratory Syncytial Virus (RSV), that are known to cause a significant burden of acute lower respiratory infection episodes in children under the age of five specifically (Shi et al. 2017). Because these first few months of life are a critical time period for immune system development, and immune responses during this time are strikingly different from the response to similar pathogens later in life (Zhang et al. 2017; Ygberg and Nilsson 2012), we set out to map the host nasal epigenome response through WGBS to respiratory viral infection in infants across ten different viruses including SARS-CoV-2. Because we will measure epigenome changes in the direct tissue of infection, this data would give us genome-wide insights into the regulatory circuitries involved in the host antiviral response, and potentially allow us to identify sources of variation leading to inter-individual differences in infection. This dataset would also allow us to identify pathways that are shared amongst groups of viruses, as well as virus-specific methylation changes. To this end, we link almost 20 million CpGs to viral response and map biological function of viral-associated CpGs within functional elements. We highlight that different respiratory viruses vary in the magnitude and direction of the alterations they cause to the host epigenome, perhaps reflecting differential immunogenicity. We note that SARS-CoV-2 and Influenza B (FluB) cause the largest magnitude of change and show similar differential methylation profiles. We show the robustness of epigenome perturbation upon SARS-CoV-2 and FluB infection through replication in independent and age-matched nasal samples. We further use single-cell RNA-sequencing as well as GWAS data to functionally validate these methylation changes and find evidence for a signature of monocyte insufficiency that may be a predisposition to infection or increased viral replication by FluB and SARS-CoV-2.

## Results

### Methylation signatures in infant nasal epigenomes upon respiratory viral infections

We generated WGBS data on 11 pools of nasal samples collected from children (<6 months) with one of the 10 respiratory viruses and age-matched non-infected infants presented at the hospital with ARI (Table 1). Where available, each sample consisted of a pool of equimolar amount of individual DNA with the following infection status and sample size: 1) N=10 Adenovirus (Adeno), 2) N=10 Coronavirus OC43 (Corona OC43), 3) N=5 Enterovirus D68 (EVD68), 4) N=10 Influenza Type A (FluA), 5) N=10 Influenza Type B (FluB), 6) N=10 Human metapneumovirus (hMPV), 7) N=10 Human Parainfluenza Virus Type 3 (PIV3), 8) N=5 Respiratory syncytial virus (RSV), 9) N=10 Rhinovirus/enterovirus (REV) and 10) N=1 SARS-CoV-2. In addition, two pools of non-infected (NI) samples were generated (Table 1).

**Table 1:** Cohort characteristics

| Virus Type | N | Mean age (months) | SD |
| --- | --- | --- | --- |
| Adeno | 10 | 3.4 | 2.2 |
| Corona Oc43 | 10 | 1.9 | 1.9 |
| EVD68 | 5 | 2.8 | 1.6 |
| FluA | 10 | 3.5 | 1.8 |
| FluB | 10 | 4.0 | 1.3 |
| hMPV | 10 | 2.8 | 1.8 |
| PIV3 | 10 | 2.4 | 1.9 |
| RSV | 5 | 2.5 | 2.3 |
| RV (REV) | 10 | 2.0 | 2.2 |
| SARS-CoV-2 | 1 | 1.0 | - |
| Negative-1 | 5 | 6.6 | 2.7 |

Each pool was sequenced at high depth (∼22.5X unique read coverage) identifying on average 25 million CpGs per pool, each at >10X (Supplementary Table 1) of which 19.2 million CpGs were covered across all samples. These data sets were then used to cluster the samples hierarchically using correlation distances (Figure 1). SARS-CoV-2 signatures appear to have the largest distance to all other respiratory viral signatures. Among the nine other signatures, FluB, FluA, and EVD68 were distinctive from the remaining six data sets, where additional sub-structure was observed in the clustering.

**Figure 1:**
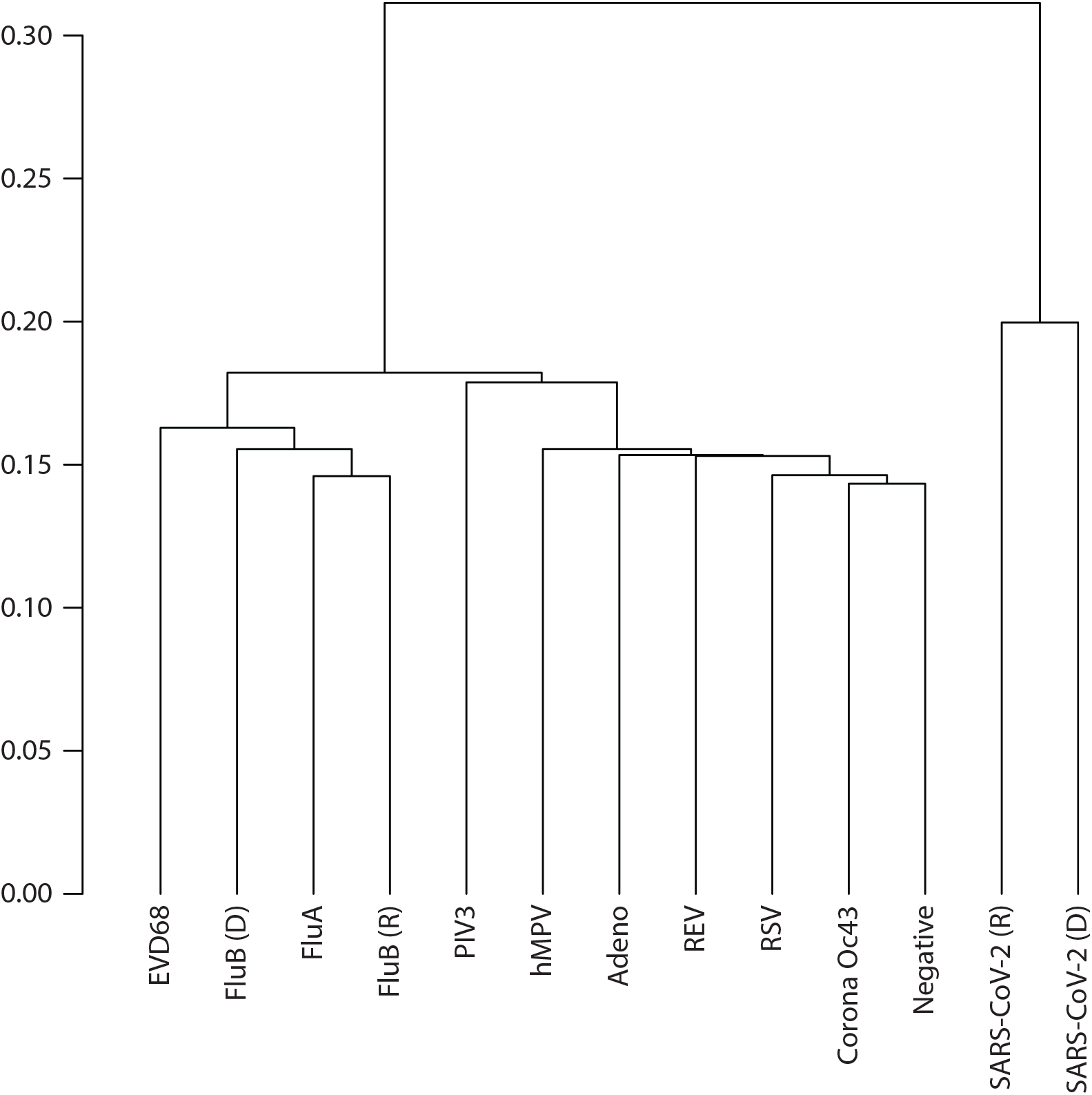
Hierarchical clustering of top 50% variable CpGs in nasal samples derived from ten respiratory viral infections.

Next, we computed genome-wide methylation differences in virus-positive vs. matched negative control samples by applying Fisher’s exact test of methylated vs unmethylated reads at CpG sites with at least 10 reads in both the pooled viral sample and the negative control. Consecutive nominally significant CpGs (p < 0.01) were grouped (N≥3) together into differentially methylated regions based on viral infection (vDMR) when having the same direction of effect and within 250bp of the adjacent CpG. For comparisons to the background nasal epigenome, we repeated the differential methylation calculations using two sets of negative control sets keeping the significance and grouping criteria the same.

Using these criteria, focusing on virus-positive vs. matched negative controls, we noted striking differences in vDMRs (Supplementary Table 2) across respiratory viruses as outlined in Figure 2A. In fact, the vDMR discovery rates follow a similar pattern as shown in the clustering analysis. Specifically, SARS-CoV-2 and FluB are associated with the strongest host response signature with SARS-CoV-2 as a clear outlier with almost 80000 vDMRs.

**Figure 2:**
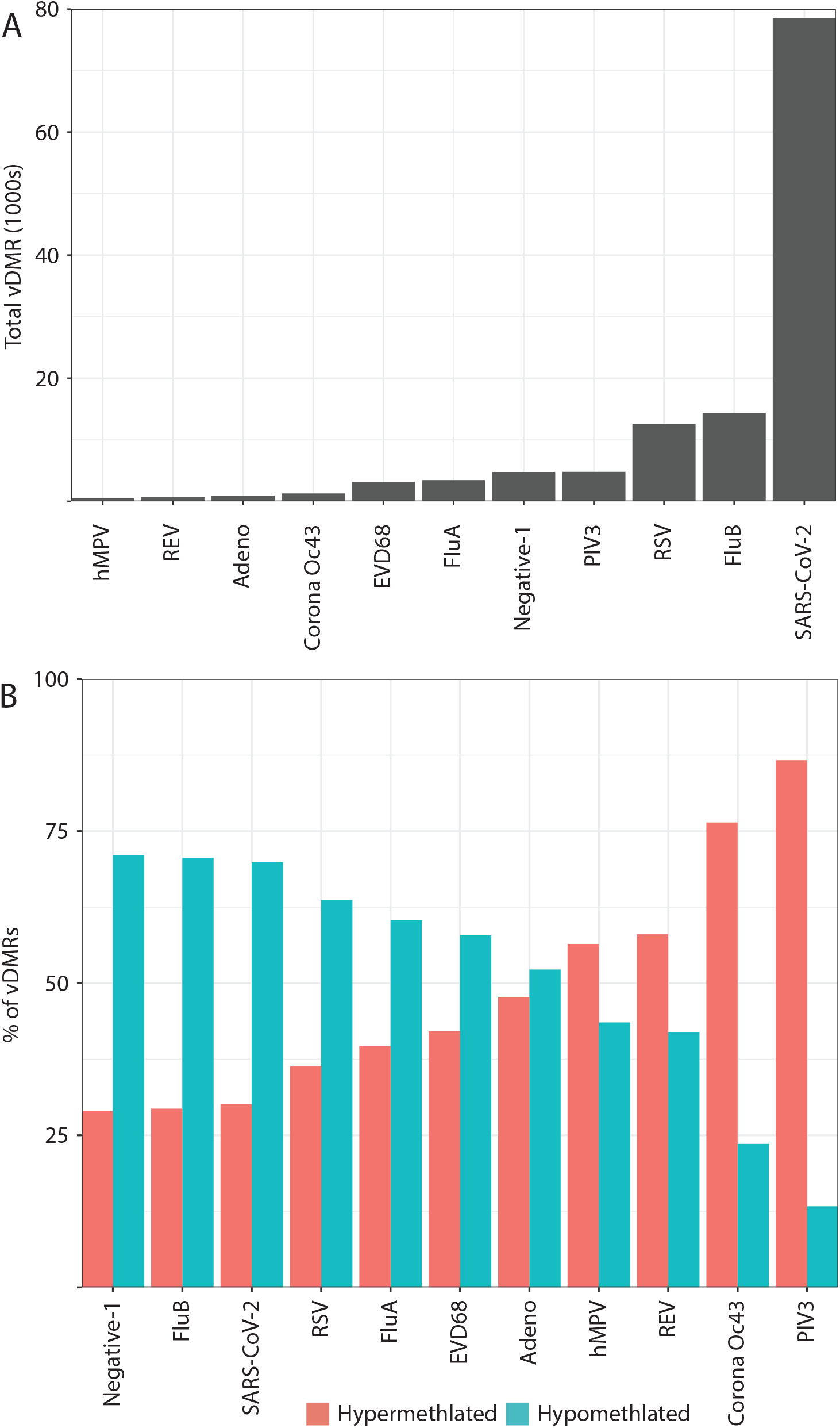
Respiratory viruses elicit differential magnitude and direction of host methylation response. A) Discovery rate of vDMR. hMPV, N=496; RV, N=665; Adeno, N=943; Corona (Oc43), N=1281; EVD68, N=3129; FluA, N=3439; PIV3, N=4788; RSV, N=12557; FluB, N=14361; and SARS-CoV-2, N=78542. B) Methylation status of vDMR

We then divided the vDMRs per virus type into hypermethylated or hypomethylated as a potential indication of deactivation/suppression vs. activation of regulatory circuitries upon viral infection, respectively. We noted that a subset of the respiratory viruses (EVD68, FluA, RSV, FluB and SARS-CoV-2) had the majority (60-70%) of their vDMRs being hypomethylated pointing towards significant activation of regulatory elements upon infection (Figure 2B). Indeed, regulatory element annotation showed that the hypomethylated vDMRs from not only EVD68, FluA, FluB and SARS-CoV-2 but also PIV3 and Adeno, were significantly more likely to map to epithelial-specific regulatory elements compared to the background control (1.5-2.5-fold, Supplementary Figure 1A) (Fisher’s Exact Test, Bonferroni p<6.25E-4). In fact, only hMPV, CoronaOC43, REV, and RSV hypo vDMR did not significantly deviate from the background control potentially indicating a milder activation of the respiratory epithelium (Supplementary Figure 1A). On the other hand, RSV hypomethylated vDMR were significantly (Fisher’s P=9.4E-15) enriched within immune-specific regulatory elements. Specifically, we found a striking overrepresentation (Fisher’s P=5.67E-20) of lymphoid-specific regulatory elements among RSV hypomethylated vDMR (Supplementary Figure 2). This observation is in line with recent evidence showing elevated levels of Type 2 respiratory innate lymphoid cells in infants with RSV infection (Norlander and Peebles 2020).

Next we annotated the hypermethylated vDMR and found in general a different pattern than for hypomethylated. For Adeno, EVD68, FluA, FluB and SARS-CoV-2, hypermethylated vDMRs were significantly enriched (Fisher’s Exact Test, Bonferroni p<6.25E-4) among immune-specific elements. This indicates deactivation or insufficiency of immune cell regulatory machineries in the host as a consequence or causing a viral infection/replication (Supplementary Figure 1B).

### Robust epigenome perturbation upon FluB and SARS-CoV-2 infection

We further evaluated the two viruses (FluB and SARS-CoV-2) causing the strongest epigenome response in the host and gathered additional age-matched samples derived from infants: 1) N=5 non-infected controls (average age 2.2 months), N=3 FluB (average age 6 months) and N=1 SARS-CoV-2 (average age 9 months). Each pool was again sequenced at high depth (∼35X unique read coverage) (Supplementary Table 1).

We first repeated the vDMR analysis (using parameters listed above) using the age-matched negative control samples and overlapped the results with the discovery FluB and SARS-CoV-2 vDMRs, respectively. We considered a vDMR to be replicated if at least one CpG per vDMR overlapped and the same direction of methylation change upon infection was observed in both discovery and replication vDMR, respectively (Supplementary Table 3).

Using this conservative threshold, we found 52% (N=7516) and 42% (N=32,318) of the vDMRs to be replicated for FluB and SARS-CoV-2, respectively. Additionally, the genome wide methylation profiles of the discovery and replicate datasets clustered together when considering the top 50% of variable CpGs, demonstrating the reproducibility of the epigenomic perturbations (Figure 1). Then, we extended the replication analysis by querying the methylation levels of all the discovery vDMRs from the replication set (N=14361 and N=78542 vDMRs for FluB and SARS-CoV-2). We noted high correlation in the SARS-CoV-2 (r=0.79) (Supplementary Figure 3A) whereas similar analysis of the FluB vDMR showed a weaker correlation across data sets (r=0.4) (Supplementary Figure 3B) supporting the notion that COVID-19 is associated with strong epigenome effects in the host.

### FluB and SARS-CoV-2 hypermethylation and lack of activation of the innate immune system

Similar to the pattern for all vDMR (Supplementary Figure 1), we found that the FluB and SARS-CoV-2 replicated hypomethylated vDMR were more likely to map to epithelial-specific regulatory elements (Figure 3A) than the background control (Fisher’s p=6.59E-29 and p=2.43E-17 for FluB and SARS-CoV-2) whereas the hypermethylated vDMRs were enriched among immune cell specific regulatory elements (Fisher’s p=1.11E-24 and p=1.72E-23 for FluB and SARS-CoV-2). We further disentangled these immune-cell specific signatures by separating regulatory elements specific to myeloid and lymphoid lineages as well as those shared across immune cells. We noted clear differences across immune cell lineages where the observed signature among hypermethylated vDMR was driven by regulatory elements specific to myeloid cells only (Figure 3B-C). In all, these results indicate a striking virus-induced activation of the epithelial-specific gene regulatory machinery but a deactivation of regulation of myeloid cells or an alternative absence of myeloid cells.

**Figure 3:**
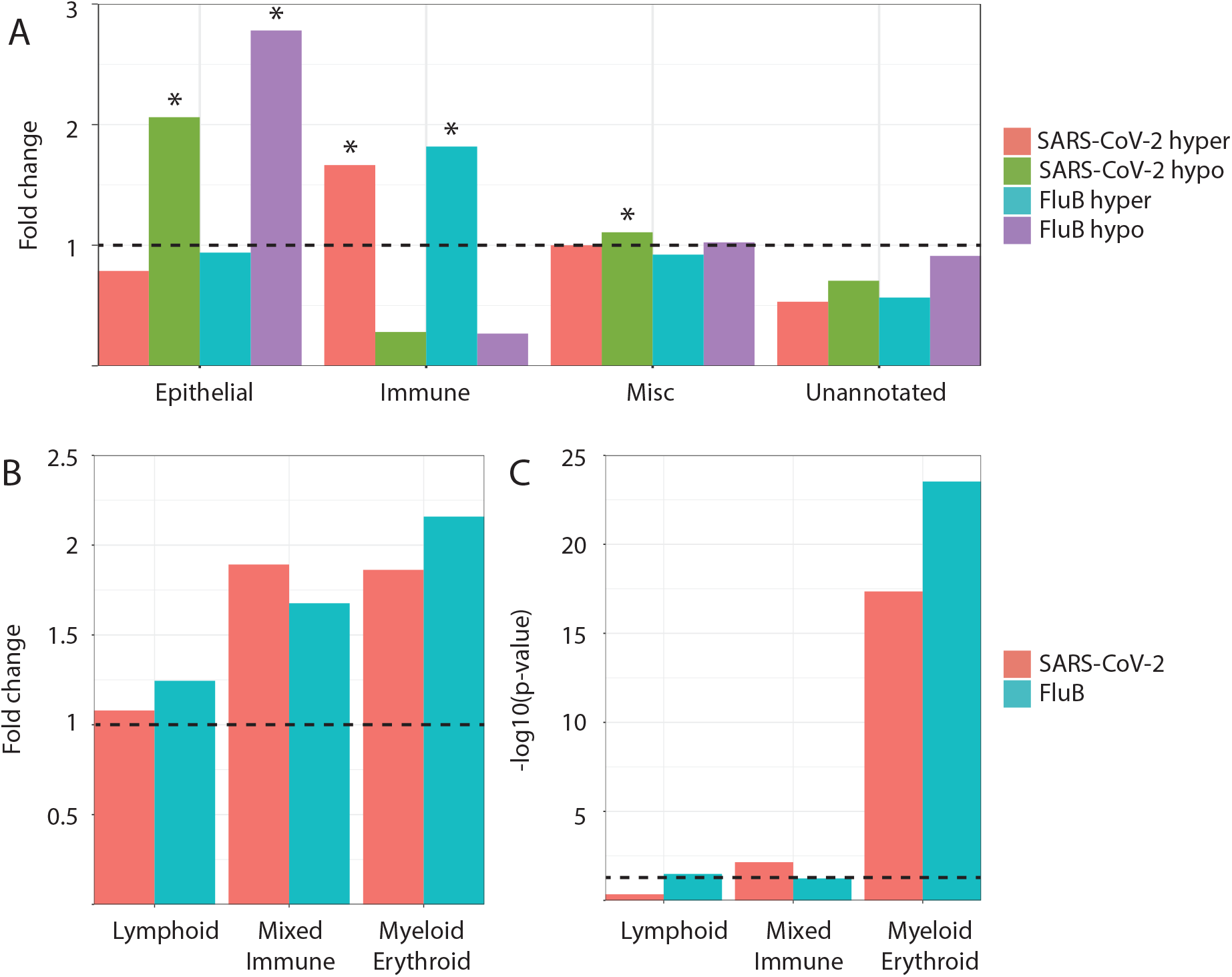
FluB and SARS-CoV-2 infection are associated with differential responses of epithelial and immune methylomes. A) Fold change compared to control sample of presence of regions annotated to specific cell lineages in replicated vDMR sets. B) Fold change compared to control sample of presence of genomic regions annotated to specific immune cell lineages in hypermethylated vDMR for FluB and SARS-CoV-2. C) -log_10_ p-value fisher’s exact test for a positive association between infection and presence of genomic regions annotated to specific immune lineages in hypermethylated vDMR. Dashed line represents p-value of 0.05.

Additionally, we queried genes associated with the vDMRs specific to FluB and SARS-CoV-2, respectively, in GREAT (5.0 kb upstream and downstream for a single nearest gene) and found leukocyte mediated immunity as the most significant biological process associated with these hypermethylated vDMRs (p=1.5E-8, Supplementary Figure 4).

Next, we used the Flu B and SARS-CoV2-specific hypermethylated vDMRs to identify potential enrichment of transcription factor binding sites (TFBS) (see Methods) to understand which regulatory pathways are associated with these viral infections. As expected, given the magnitude of vDMRs in SARS-CoV-2 vs. FluB comparisons, we found almost twice as many enrichments in SARS-CoV-2 vs FluB hyper vDMR at nominal p < 0.01 (N=145 vs N=74, Supplementary Table 4-5). We performed Gene Ontology (GO) term enrichment analysis of TFBS enrichments at P<10^−10^ which corresponded to 50 and 22 unique TF genes for SARS-CoV-2 and FluB, respectively, with a 100% overlap of FluB TF genes among the SARS-CoV-2 genes. Among these GO biological processes annotations, myeloid cell differentiation was the top enriched term (P=5.12E-13, Supplementary Table 6) potentially indicating absence of sufficient activation of the innate immune system and thus increased viral replication. In fact, the top enriched TFBS for both SARS-CoV-2 and FluB hyper vDMR corresponded to *ELF4 (*p=1E-208 and p=1E-51; Supplementary Table 4-6) which is known to be critical for antiviral immunity and host defense by activating innate immunity (You et al. 2013; Szabo and Rajnavolgyi 2014). We examined the expression pattern of *ELF4* across hematopoietic cells using publicly available data (Stunnenberg et al. 2016) and found highest expression in cells of the myeloid lineage (Supplementary Figure 5).

### Functional analysis of replicated vDMR in childhood infection

To investigate the functional roles of the replicated vDMRs for both FluB and SARS-CoV-2, we performed single-cell RNA-sequencing (scRNAseq) on pooled nasal mucosal samples from children infected with Flu B (n=4, average age = 6.25 years), SARS-CoV-2 (n=3, average age = 7.66 years), and from uninfected and age-matched controls (n=5, average age = 7 years). We captured and sequenced over 12,000 cells across the pools (FluB = 3,687 cells, SARS-CoV-2 = 2,864 cells, control = 4,963 cells), and were able to identify all expected major cell types (Figure 4A).

**Figure 4:**
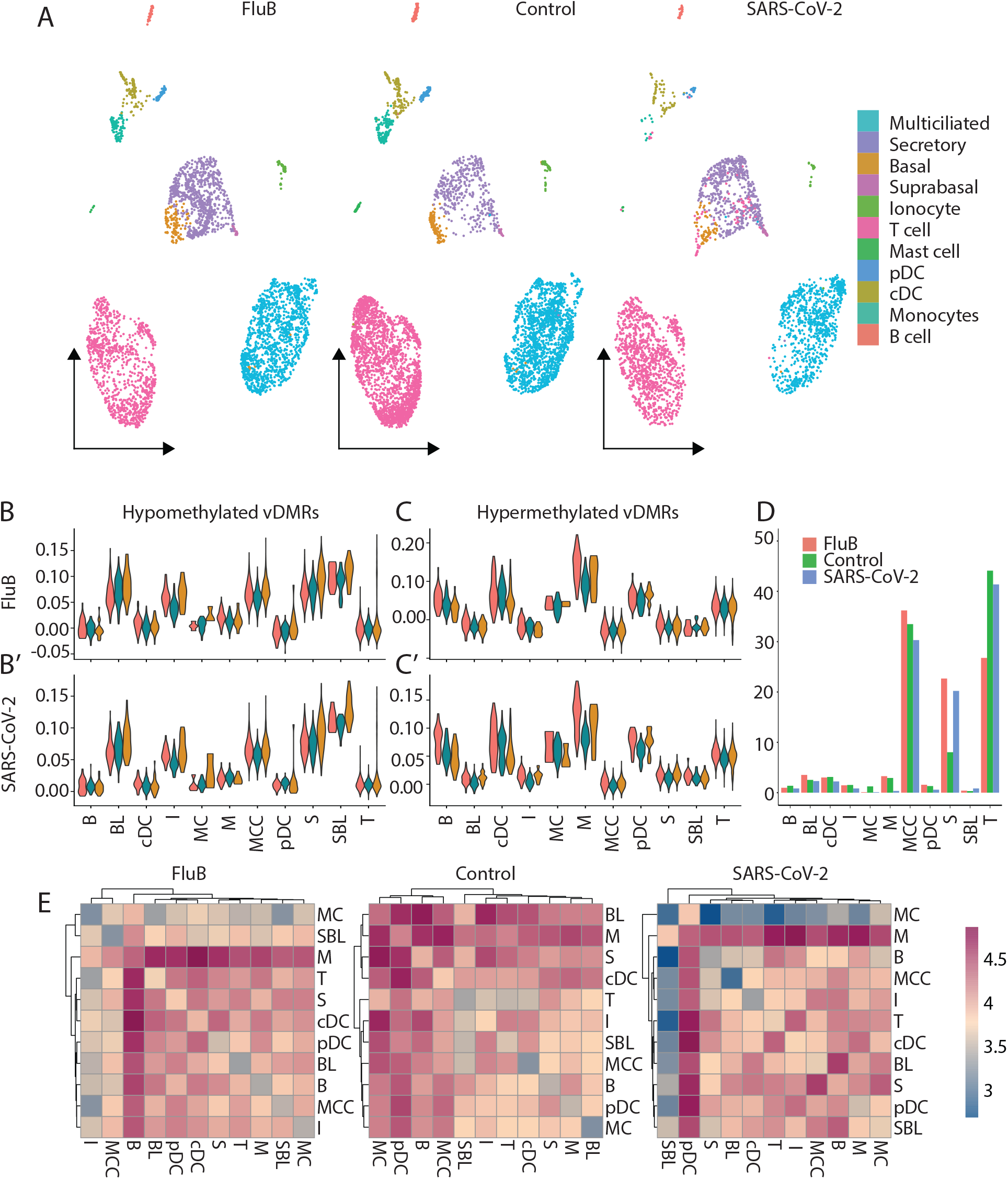
Functional analysis of replicated vDMR in adolescent infection. A) UMAP projection of cells obtained from pooled adolescent nasal mucosa samples infected with FluB, SARS-CoV-2 and uninfected age-matched controls. B-C) Expression level of genes “modules” derived from genes nearest vDMR for Flu B (B) and SARS-CoV-2 (C). D) Percent of total cells in each cell type for uninfected and infected pooled samples. E) Heatmaps of cell-cell interactions between all cell types for FluB infected, SARS-CoV-2 infected, and uninfected controls.

We examined the expression level of gene “modules” consisting of those genes within 5 kb of a replicated vDMR for FluB and SARS-CoV-2. First, we observed that the gene modules associated with hypomethylated vDMRs for FluB and SARS-CoV-2 are more highly expressed in epithelial cell types than in immune cell types (Figure 4B-B’). Interestingly, the gene modules associated with hypermethylated vDMRs for FluB and SARS-CoV-2 are more highly expressed in immune cell types than epithelial cell types, thus corroborating our previous cellular annotations of vDMRs (Figure 4C-C’). In fact, we observed the largest increase of expression of these genes after infection in monocytes (Figure 4C).

In an attempt to clarify if the observed hypermethylation of myeloid associated genomic regions was a consequence of infection or represented a potential absence of myeloid cells which functions as an epigenetic predisposition to infection or increased viral replication, we examined the activity of myeloid cells after infection in both FluB as well as SARS-CoV-2. We hypothesized that if the observed hypermethylation of myeloid regulatory genomic regions was a consequence of viral infection, we should see a decrease in either cell number or activity of myeloid cells compared to controls. We found that the proportion of myeloid cells was not fundamentally changed after either viral infection compared to controls (Figure 4D). To assess myeloid cell activity after infection, we quantified the number of cell-cell interactions through ligand receptor pairs. We found that monocytes had the largest number of interactions with other cell types after both FluB and SARS-CoV-2 infection (Figure 4E) and were consistently increased in their cell-cell interactions compared to controls after infection in both SARS-CoV-2 and FluB (Supplementary Figure 6). These results together indicate that myeloid cell activity and proportion in the nasal mucosa is not decreased as a result of FluB or SARS-CoV-2 infection, but rather an environmental or genetic predisposition to a viral infection and replication due to insufficient resident myeloid cells in the nasal mucosa may be the cause for the observed hypermethylation of myeloid regulatory regions as opposed to active immune suppression by viral infection.

### Virus-associated hypermethylation overlap disease loci

To test the hypothesis of a potential genetic predisposition, we used genetic loci identified in large-scale genome-wide association studies (GWAS) to guide our interpretation of the biological consequences of these environmentally perturbed epigenome variations. With the observed enrichment of myeloid immune cellular pathways among the hypermethylated pattern in regulatory elements associated with both FluB and SARS-CoV-2, and the lack of functional evidence for myeloid cell suppression in the scRNAseq data, we naturally opted to utilize genetic information linked to human blood cell trait variation identified from a large meta-analysis of GWAS to test our hypothesis that resident myeloid cell absence may be a predisposition for FluB and SARS-CoV-2 infection. Specifically, we integrated common genetic loci associated with hematological traits from the largest GWAS to date identifying almost 17000 genetic variants across 28 blood cell phenotypes (Vuckovic et al. 2020). We assessed the co-localization of these variants within the 7516 and 32318 Flu B and SARS-CoV-2 vDMR and contrasted similar co-localization pattern within randomly selected control regions matching the genomic contexts of the vDMR (N=49623). We performed Fisher’s Exact test to evaluate the difference in proportional overlap in test (vDMRs) vs. control regions, respectively. Intriguingly, we found monocyte proportion as the most significant trait both for hypermethylated FluB (Fisher P=4.85E-5) and hypermethlyated SARS-CoV-2 vDMR (Fisher P 1.06E-8), respectively (Supplementary Figure 7, Supplementary Table 7)), indicating that observed interindividual variation in susceptibility to viral infections may have a genetic basis.

## Discussion

We have demonstrated the breadth of host epigenomic alterations occurring in infants in response to 10 common respiratory viruses during childhood including SARS-CoV-2. We find that while there is substantial variability in the magnitude of methylome response to different viral infections, many of the identified DMRs are reproducibly identifiable between independent infections, indicating robust epigenome response to infection. Additionally, we demonstrate an important role for innate cells, specifically monocytes, in the antiviral response to influenza B and SARS-CoV-2.

While our functional analysis of vDMR for FluB and SARS-CoV-2 through scRNAseq confirms that genes near hypomethylated vDMR do increase in expression after infection, the converse was not true for hypermethylated vDMRs. Typically, hypermethylation is interpreted as a repression of the specific genomic region involved. However, an alternative explanation is that a heterogenous cell population contains fewer cells of the lineage associated with a specific regulatory region. Given that genes near hypermethylated vDMRs for FluB and SARS-CoV-2 actually increase in expression after infection, that these hypermethylated vDMRs are enriched for regions that are associated with monocyte development, and that monocyte activity increases in terms of cell-cell interactions after infection, it is unlikely that viral infection with FluB or SARS-CoV-2 causes hypermethylation of these regions. Instead, we propose that these hypermethylated regions might represent a predisposition to viral infection or viral replication by indicating an absence or reduced presence of monocytes. This would require further testing in single individuals to directly measure methylation status of vDMRs before and after infection, however the importance of myeloid cells and specifically monocytes in response to these viral infections among infants is clear.

## Conclusions

In this work, we establish a catalogue of the methylation response to respiratory viral infection in infants. We find differences in the epigenomic response to different viruses as well as differential responses across the age range. Finally, we demonstrate that an epigenomic signature of monocyte suppression may actually reflect predisposition to infection or viral replication and may account for inter-individual differences in infection propensity and immune response.

## Methods

### WGBS Sample characteristics

Salvage nasal mucosa derived from children presenting with an acute respiratory illness at Children’s Mercy were accessed and collected from pediatric mid-turbinate nasal flocked swabs as part of routine testing for pathogens. Samples were stored in 3ml of Universal Transport Medium where 200ul of each specimen was tested by BioFire respiratory pathogen panel or for SARS-CoV-2 and remaining aliquot was saved in −80C freezer. In the current study and prior to March 2020, samples derived from infants less than 6 months of age were selected across the following groups: Adenovirus positive, Coronavirus OC43 positive, Enterovirus D68 positive, Influenza Type A positive, Influenza Type B, Human metapneumovirus positive, Human Parainfluenza Virus Type 3 positive, Respiratory syncytial virus positive, Rhinovirus positive and Pathogen Panel Negative, respectively. From March 2020, samples were also derived from SARS-CoV-2 positive infants. An additional set of SARS-CoV-2 (20 months and 19 years) and Influenza Type B positive samples (0-6 months, 6-12 months, 12-24 months, 2-5 years) were selected across different age-groups.

### Specimen Pooling and DNA Isolation

Nasal specimens were stored at −80°C and were brought to room temperature before pooling by viral type or age-group if applicable. Before pooling, the specimens were mixed well with gentle pipetting. In total of 100µL from each specimen of the same viral type or age group was removed and pooled together in a 1.5mL tube. Once all specimen aliquots were added to the viral pool, the pool was mixed by pipetting and 200μl was taken from each pool into a new 1.5mL tube for DNA isolation. Similarly, single samples (SARS-CoV-2 positive) was mixed by pipetting and 200μl was taken from each sample into a new 1.5mL tube for DNA isolation. DNA was isolated with a DNeasy Blood and Tissue Kit (Qiagen, Cat No. 69504) with the following modifications to kit protocol: 8uL of RNase A was used instead of 4ul during the optional RNase A step and the lysis incubation time at 56°C was increased to at least 3 hours to ensure complete lysis of the specimens. After isolation, the DNA concentration of each sample was determined using a Qubit dsDNA HS Assay Kit (Fisher, Cat No. Q32851).

### WGBS Library Preparation and Sequencing

In total of 100ng of DNA was aliquoted from each sample pool. Unmethylated λDNA was added to each sample at 0.5%w/v and the samples were sheared mechanically using a Covaris LE220-plus system to a length of 350 bp, using the settings recommended by the manufacturer. The sizing was determined by a High Sensitivity D1000 ScreenTape and Reagents (Agilent, Cat. No. 5067-5584 and 5067-5585) on the TapeStation platform. Once the input DNA was at the proper fragment size, the samples were concentrated with a SpeedVac to a volume of 20µL. The samples then underwent bisulfite conversion with an EZ DNA Methylation-Gold kit (Zymo, Cat. No. D5006). The samples were eluted off the spin columns with 15μl of low EDTA TE buffer (Swift, Cat. No. 30024) before library preparation.

The low-input libraries were prepared using an ACCEL-NGS Methyl-Seq Library kit (Swift, Cat. No. 30024) with a Methyl-Seq Set A Indexing Kit (Swift, Cat. No. 36024), following the protocol associated with the library kit. During the protocol, bead cleanup steps were performed with SPRIselect beads (Beckman Coulter, Cat. No. B23318). Following the recommendation of the kit, 6 PCR cycles were performed to amplify the samples. The final libraries were quantified with a Qubit dsDNA HS Assay Kit and the size was determined by using a BioAnalyzer High Sensitivity DNA Kit (Agilent, Cat. No. 5067-4626). The libraries were then sequenced on the Illumina NovaSeq6000 System using 150bp paired-end sequencing.

### WGBS data processing

WGBS data was processed using the Epigenome Pipeline available from the DRAGEN Bio-IT platform (Edico Genomics/Illumina). Sequence reads were demultiplexed into FASTQ files using Illumina’s bcl2Fastq2-2.19.1 software and trimmed for quality (phred33 >= 20) and Illumina adapters using trimgalore v.0.4.2 (https://github.com/FelixKrueger/TrimGalore). Reads were then aligned to the bisulfite-converted GRCh37 reference genome using DRAGEN EP v2.6.3 in paired-end mode using the directional/Lister methylation protocol presets. Alignments were calculated for both Watson and Crick strands and the highest quality unique alignment was retained. Duplicated reads were removed using picard v 2.17.8 (Broad Institute 2019). A genome-wide cytosine methylation report was generated by DRAGEN to record counts of methylated and unmethylated cytosines at each cytosine position in the genome. Methylation counts were provided for the CpG, CHG and CHH cytosine contexts but only CpG was considered in the study. To avoid potential biases in downstream analyses, CpGs were further filtered by removing CpGs: covered by five or less reads, and located within genomic regions that are known to have anomalous, unstructured, high signal/read counts as reported in DAC blacklisted regions (DBRs) or Duke excluded regions (DERs) generated by the ENCODE project (Amemiya et al. 2019).

### Differential Methylation Analysis

Filtered methylation data from all nasal samples derived from infants were merged, and only CpGs covered by at least 10 reads were kept. The Fisher’s exact test for a 2×2 contingency table was used to evaluate the difference in methylated vs. unmethylated reads in the viral sample compared to the matching negative control at each CpG sites. Consecutive nominally significant CpGs (p < 0.01) were grouped together into blocks when having the same direction of effect and within 250bp of the adjacent CpG and only region with three or more CpGs were kept for further analysis.

### Annotation of regulatory elements

DNase I Hypersensitive Site (DHS) coordinates were accessed from https://zenodo.org/record/3838751#.YEY9qRBKhTZ using 16 different vocabulary representatives as outlined in (Meuleman et al. 2020)

### Transcription Factor Binding Analysis

Transcription factor binding site (TFBS) motif analysis was performed using the Homer software (HOMER findMotifsGenome.pl v4.11.1) (Burger et al. 2013) using the central 200bp of regions. The UMRs and LMRs called from the merged superset sample of all viral pools were used the background.

### scRNA-seq Patient Recruitment

All study subjects were enrolled at Children’s Mercy either using salvage sample collection protocol or using prospective cohort study protocols. Specifically, the NM cohort of control individuals included patients tested for COVID-19 as a part of their standard of care procedure, these samples were collected using a salvage sample protocol (IRB # STUDY00001258). Similarly, patients undergoing multiplex testing for respiratory viruses were regularly screened and all children positive for Influenza B were selected for the NM cohort of Influenza B positive children these samples were collected using salvage sample protocol (IRB # STUDY00001193). For COVID-19 positive children, families were enrolled in the CODIEFY study approved by the Institute Review Board (IRB) at Children’s Mercy (IRB # STUDY00001317). Parents or legally appointed representatives of COVID-19 positive children were approached for enrollment and verbal consent within 24-48 hours of their test results, and children aged 7 years and above have given verbal informed assent. Respiratory specimens were collected and transported by a home-health care nurse following standard precautions within the next 24-48 hours. Samples were processed for nasal cell isolation within 2 - 4 hours of collection.

### Single-cell RNA-sequencing

Samples were collected from pediatric mid-turbinate nasal flocked swabs and were stored in 3ml of Universal Transport Medium where 200ul of each specimen was tested by BioFire respiratory pathogen panel or for SARS-CoV-2 and remaining aliquot was kept in 4C until test result was available (within 12h). 1ml of each sample was diluted with cold PBS (Thermo Fisher Cat No. 14190144) + 2% FBS (GE Healthcare Cat No. SH30088.03HI) up to a total volume of 5 mL and passed through a 40-µm nylon mesh cell strainer that had been prewetted with 2 mL of PBS + 2% FBS. The strainer was then rinsed with 7 mL of cold PBS + 2% FBS. The sample was transferred to a 15-mL conical tube and centrifuged at 300 x g at 4°C for 8 minutes. The supernatant was carefully removed without disturbing the cell pellet. The cell pellet was resuspended in 200 µL of cold PBS + 2% FBS, and the cell count and viability were assessed using 0.4% Trypan Blue and a Countess II automated cell counter. The cell suspension was transferred to a 1.5-mL tube and centrifuged at 300 x g at 4°C for 8 minutes, and the supernatant was carefully removed without disturbing the cell pellet. The cell pellet was resuspended in 1 mL of cold Recovery Cell Culture Freezing Medium (Thermo Fisher Cat No. 12648010), and the cell suspension was transferred to a cryogenic storage vial. The cryogenic storage vial was placed in a Corning CoolCell FTS30, which was then placed in a −80°C freezer overnight. Samples were stored at −80°C for no longer than one week before being thawed and processed for scRNAseq. Upon thawing, sample with less than 30% viability were excluded from analysis and cells were used in pools or individually. For the Influenza B positive samples, a single pool of four samples was created (4-11 years, n=4). COVID-19 positive samples (5-11 years, n=3) were processed individually. For each sample to be thawed, 10 mL of Thawing Medium consisting of DMEM/F-12 (Thermo Fisher Cat No. 11320033) supplemented with 10% FBS and 100 units/mL of penicillin and 100 µg/mL of streptomycin (Thermo Fisher Cat No. 15140122) was prewarmed in a 37°C bead bath. Each cryogenic storage vial containing a sample to be thawed was placed in the 37°C bead bath. No more than 5 samples were thawed at a time. When only a small ice crystal remained in the sample, both the cryogenic storage vial and the 15-mL conical tube containing the Thawing Medium were aseptically transferred to the biosafety cabinet. 1 mL of Thawing Medium was slowly added, dropwise, to the sample. The diluted sample was then mixed gently by pipetting and further diluted in the remaining 9 mL of

Thawing Medium. The thawed and diluted cells were left at room temperature while the remaining samples were similarly thawed. When all samples in the batch were thawed, the samples were centrifuged at 300 x *g* for 8 min. The supernatant was carefully removed without disturbing the cell pellets. The cell pellets were each resuspended in 0.5 mL of Thawing Medium, and the cell suspensions were placed on ice. Each pool or individual sample was passed through a prewetted 40-µm nylon mesh cell strainer, and the cell strainers were rinsed with 5 mL of cold Thawing Medium. The pooled or individual sample cell suspensions were centrifuged at 300 x *g* for 8 min at 4°C, and the supernatant was carefully aspirated without disturbing the cell pellets. The cell pellets were resuspended in 100 µL of cold Thawing Medium, and cell count and viability were assessed using 0.4% Trypan Blue and a Countess II automated cell counter. For the Influenza B group, 2 wells of a Chromium Chip B (10x Genomics Cat No. 1000153) were loaded with 32,000 cells each; for each SARS-CoV-2 positive sample, 2 wells of a Chromium Chip B (10x Genomics Cat No. 1000153) were loaded with 16,000 cells each. Following cell loading, scRNAseq was performed identically for all samples using the Chromium Single Cell 3’ Library & Gel Bead Kit v3 (10x Genomics Cat No. 1000075) according to the manufacturer’s protocol. Sequencing was performed using an Illumina NovaSeq 6000. Runs of WGBS were 2×151 cycle paired-end, while runs of scRNAseq were 2×94 cycle paired-end.

### Post-sequencing analysis scRNAseq

Sequenced reads were initially processed by the cellranger pipeline (v3.1.0) which includes fastq creation, read alignment, gene counting, and cell calling. All samples were mapped to the cellranger GRCh38 v1.2.0 genome. The resulting cell by gene matrix from the cellranger “count” step was then processed using standard workflows in Seurat (Butler et al. 2018; Stuart et al. 2019). In brief, low quality cells were removed by filtering out cells with a unique gene count lower than 750 and more than 50% mitochondrial reads. The gene counts for remaining cells that passed quality control were then normalized using SCTransform (Hafemeister and Satija 2019) with the replicate captures as a batch variable. The COVID-19 positive and Influenza B positive samples, were normalized independently, and integrated with the control samples using the FindIntegrationAnchors and IntegratedData functions in Seurat with default parameters. The integrated data was then used for linear and non-dimensional reduction, nearest neighbor finding, and unsupervised clustering. Cell types were assigned by examining expression of known genes in the unsupervised clusters, as well as examining markers of the clusters identified using the FindAllMarkers function in Seurat with default parameters.

### Cell-cell interaction quantification

We quantified the interactions between different cell types in the nasal epithelia of pediatric patients using the cellphoneDB program (Efremova et al. 2020). The SCTransform corrected gene expression values for each infection state were independently input into cellphoneDB using the “statistical analysis” pipeline. The resulting cell-type by cell-type matrix of statistically significant interactions for each infection state were then plotted as clustered heatmaps, using a consistent color scale between the different infection states.

### Gene expression module scoring

Gene expression modules for Interferon stimulated genes and cell death markers were scored using the AddModuleScore function in Seurat. The genes used for each of these modules can be found in Supplementary Table 8.

## Data Availability

All raw and processed sequencing data generated in this study have been submitted to the NCBI Gene Expression Omnibus (GEO; https://www.ncbi.nlm.nih.gov/geo/) under accession number GSE168254 and GSE162864. Fully processed single-cell data are available for exploration through the UCSC cell browser (lifespan-nasal-atlas.cells.ucsc.edu)

https://lifespan-nasal-atlas.cells.ucsc.edu

## Abbreviations

SARS-CoV-2: Severe acute respiratory syndrome – Coronavirus – 2
FluB: Influenza Type B
FluA: Influenza Type A
DMR: differentially methylated region
vDMR: viral differentially methylated region
ARI: acute respiratory illness
WGBS: whole genome bisulfite sequencing
TFBS: Transcription factor binding site
Adeno: Adenovirus
Corona: Oc43
EVD68: Enterovirus D68
hMPV: Human metapneumovirus
PIV3: Human Parainfluenza Virus Type 3
RSV: Respiratory syncytial virus
REV: Rhino(entro)virus
GWAS: Genome-wide association study
scRNAseq: single-cell RNA-sequencing

## Declarations

### Ethics approval and consent to participate

The complete WGBS project and single-cell profiling using pooled samples was determined as non-human subjects research by the Institutional Review Board (IRB) at Children’s Mercy Research Institute. Single sample single-cell profiling (SARS-CoV-2) study was approved by the IRB (STUDY00001317) at Children’s Mercy Research Institute.

### Consent for publication

All study participants and their family members enrolled in human subjects’ research (i.e. single-cell profiling of SARS-CoV-2 positive nasal samples) provided informed consent for publication.

### Authors’ contributions

EG, RS, and TP conceived the study. KW, BK, and WC analyzed data. RS and DB provided samples. KW, and EG prepared the manuscript with significant contribution by TP and RS. All authors read and approved the manuscript.

## Acknowledgments

We thank Bradley Belden, Rebecca Biswell, Daniel Louiselle, Nyshele Posey and Margaret Gibson at the Genomic Medicine Center at Children’s Mercy Kansas City for technical assistance and clinical coordination.

## Figure legends

**Supplementary Figure 1:**
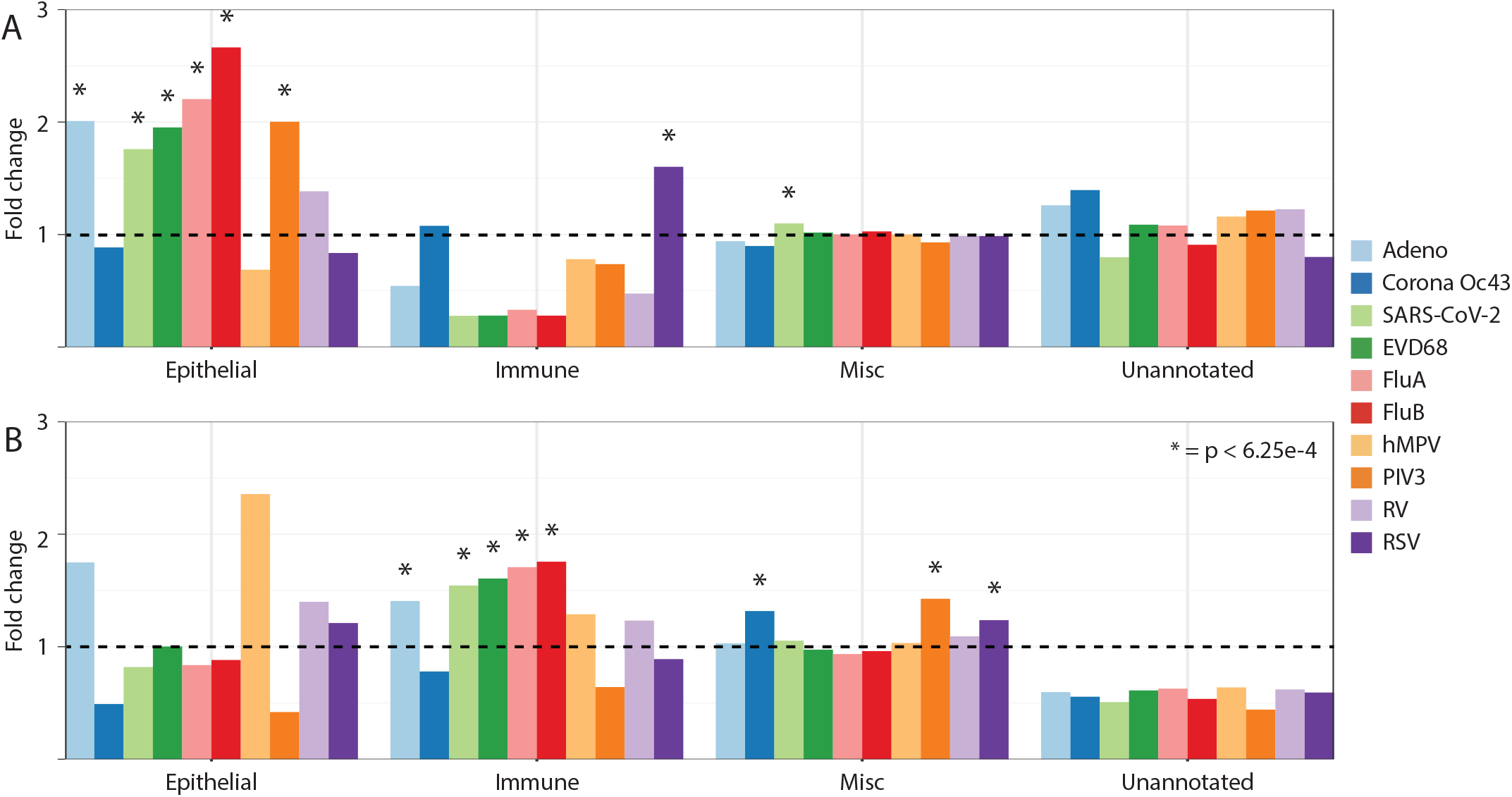
Enrichment of regulatory element annotations of hyper (A) and hypo (B) vDMR. Fold change shown is compared to control sample.

**Supplementary Figure 2:**
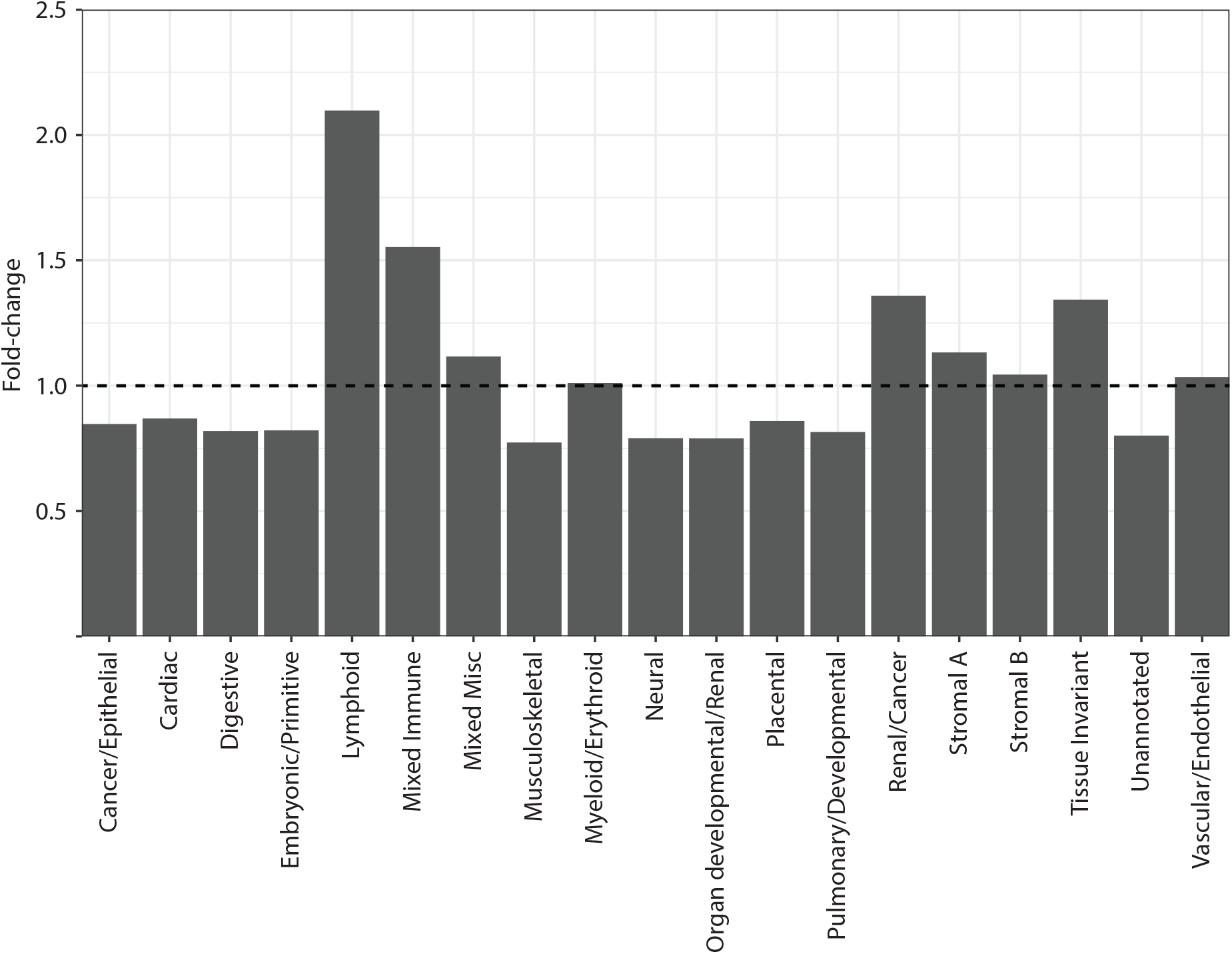
Enrichment of regulatory element annotations of RSV hypo vDMR. Fold change shown is compared to control sample.

**Supplementary Figure 3:**
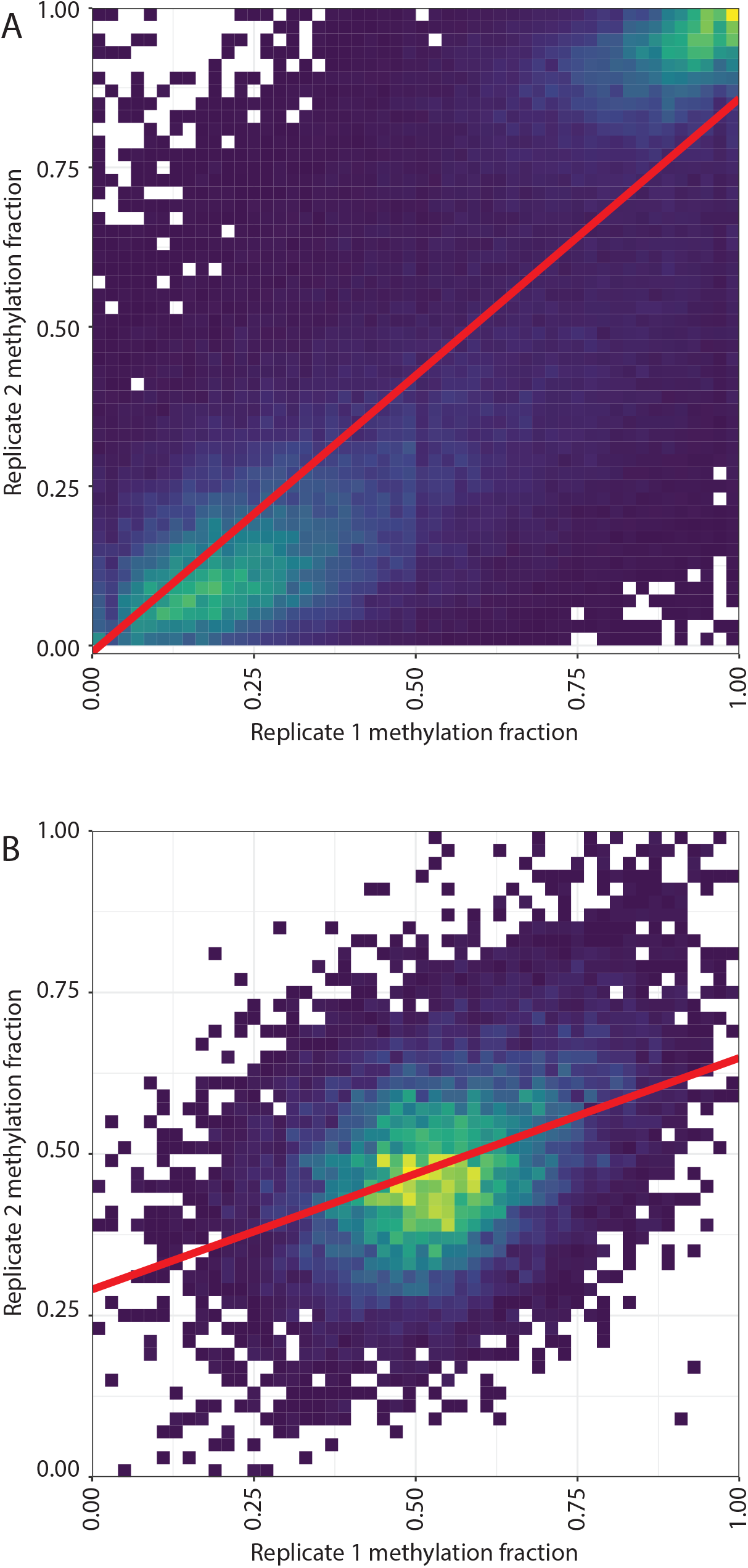
Density plots of methylation level across discovery and replication set for SARS-CoV-2 (A) and Flu B (B).

**Supplementary Figure 4:**
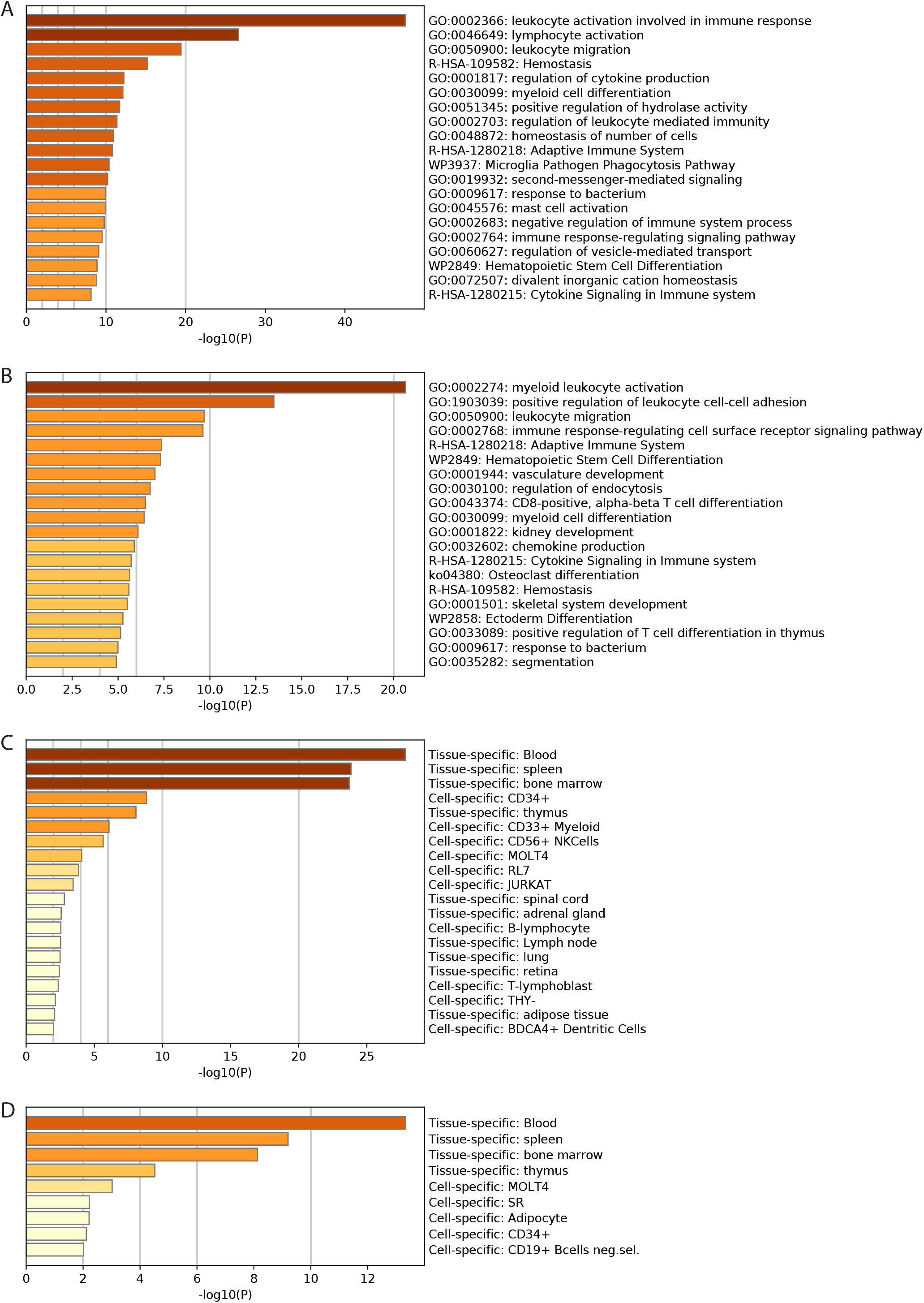
FluB and SARS-CoV-2 hypermethylated vDMR are nearest genes that are enriched for innate immune cell development terms. A-B) GO term enrichment for genes that are nearest hypermethylated vDMRs for SARS-CoV-2 (A) and FluB (B). C-D) Expression domain enrichment for genes that are nearest hypermethylated vDMRs for SARS-CoV-2 (C) and FluB (D).

**Supplementary Figure 5:**
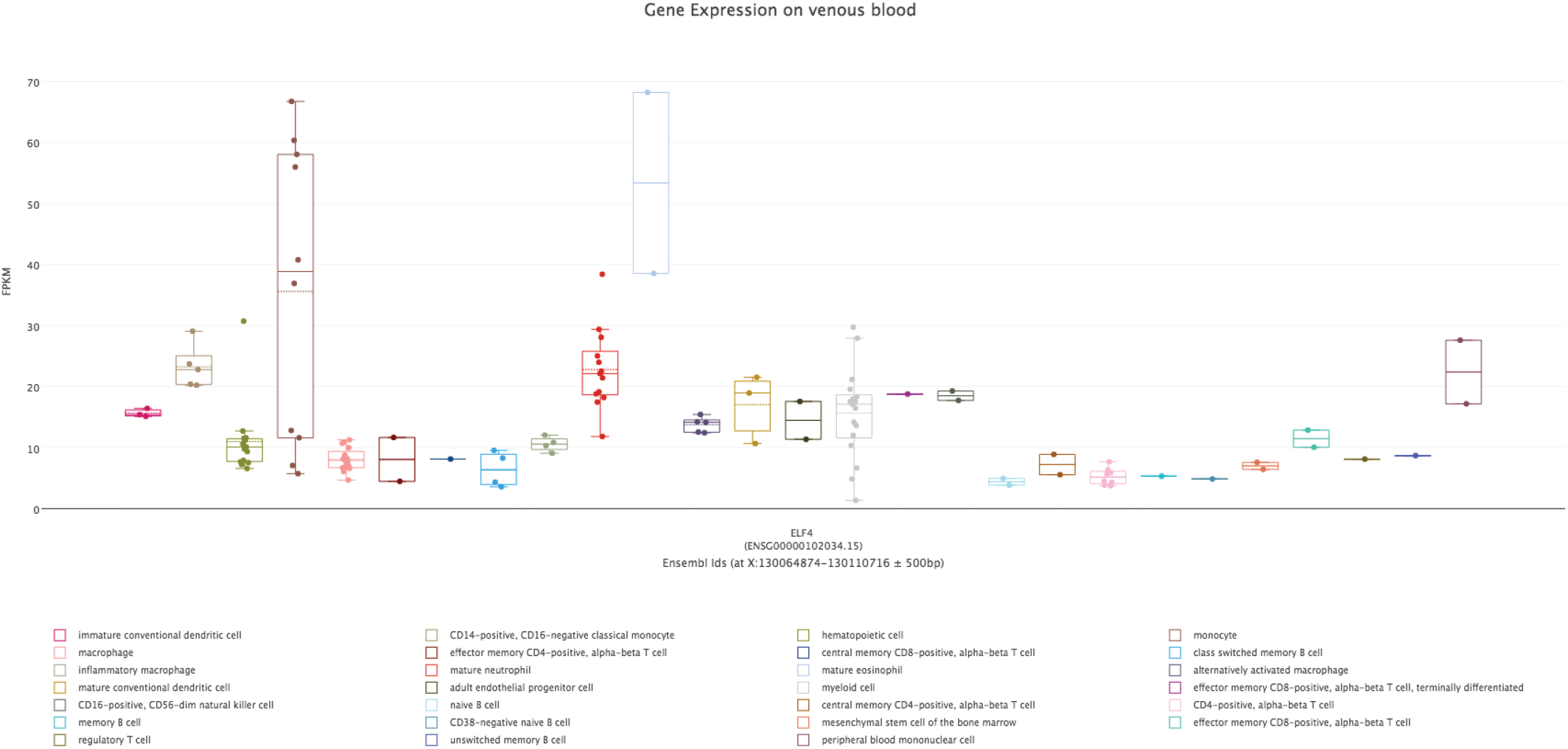
Expression of *ELF4* in different cell lineages present in venous blood demonstrates enrichment in monocytes.

**Supplementary Figure 6:**
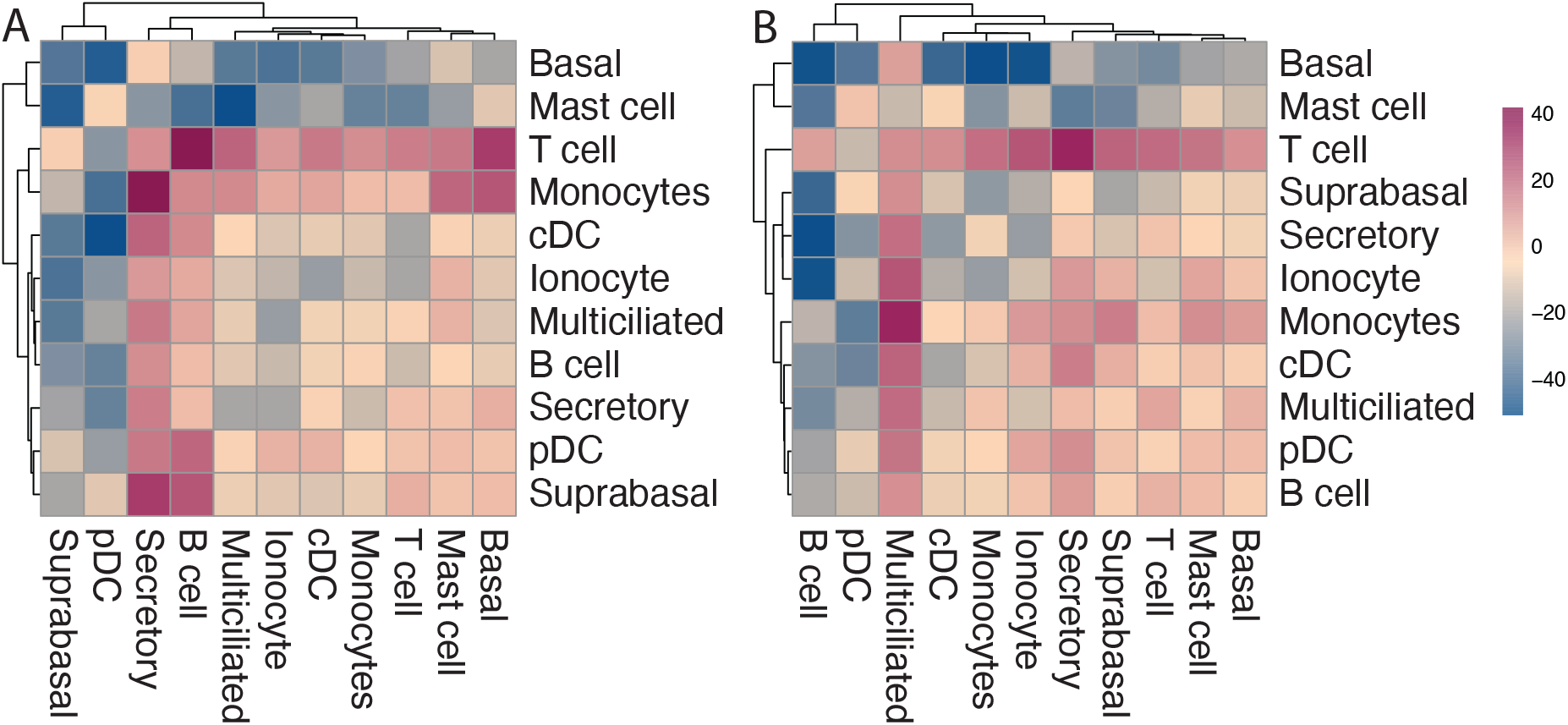
Genomic regions associated with monocyte development determined by GWAS are the most significantly enriched regions in hypermethylated vDMR for FluB and SARS-CoV-2.

**Supplementary Figure 7:**
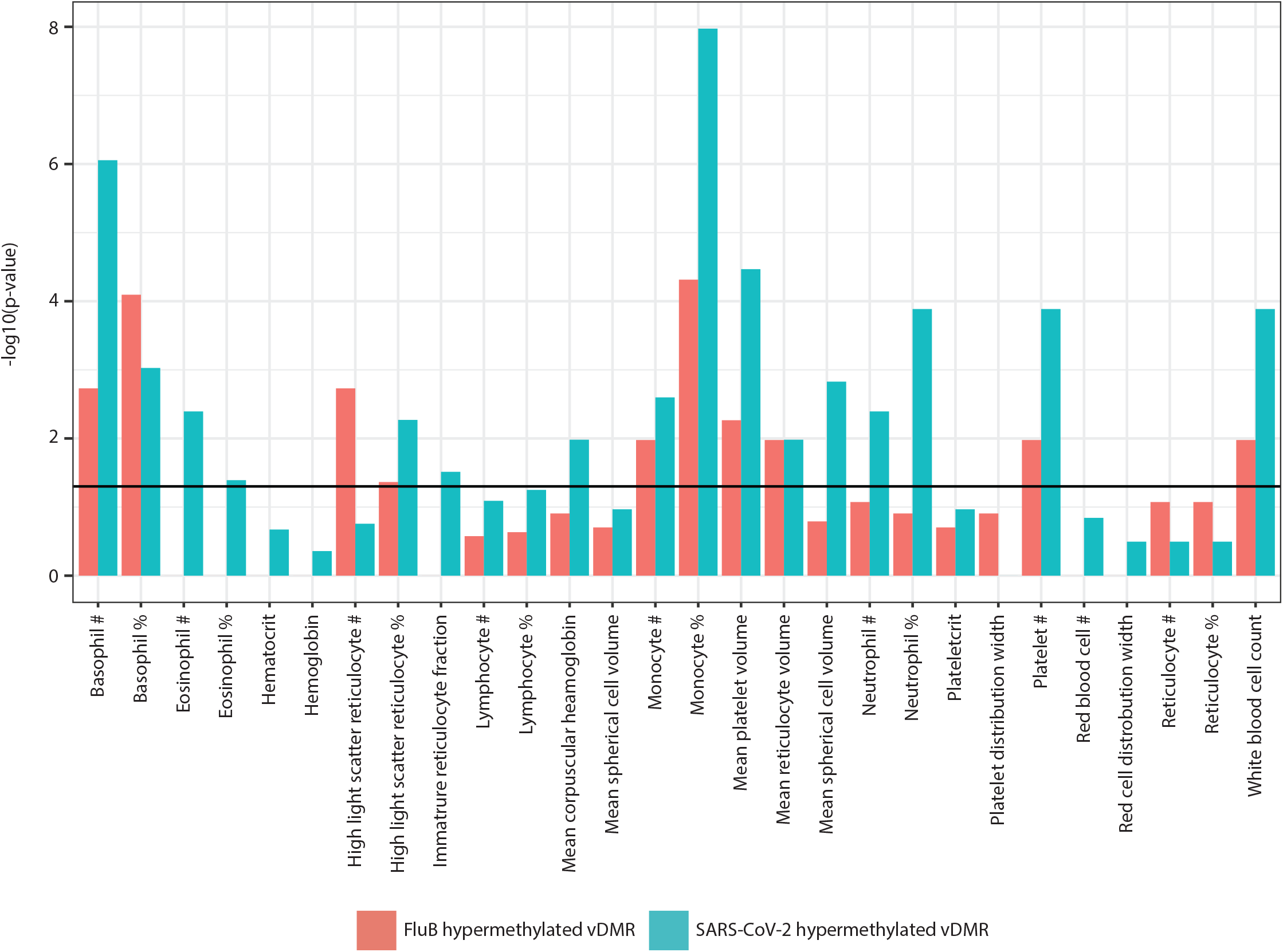
Monocytes repeatably increase cell-cell interactions after infection. A-B) Change in number of interactions between each cell-type pair after infection with SARS-CoV-2 (A), and Flu B (B). Solid line represents a p-value of 0.05.

